# Cold pressor test and paradoxical blood pressure reduction in AL amyloidosis

**DOI:** 10.1101/2025.03.10.25323710

**Authors:** Raphael Patras, Georgios Georgiopoulos, Foteini Theodorakakou, Ioannis Petropoulos, Dimitris Delialis, Lasthenis Angelidakis, Alexandros Briasoulis, Maria Gavriatopoulou, Panagiotis Kokotis, Efstathios Manios, Meletios-Athanasios Dimopoulos, Efstathios Kastritis, Kimon Stamatelopoulos

**Author notes:** Address for Correspondence: Prof. Kimon Stamatelopoulos, MD, Vascular Laboratory, Department of Clinical Therapeutics, Alexandra Hospital, Medical School, National and Kapodistrian University of Athens, PO Box 11528, 80 Vas. Sofias Str., Athens, Greece. & Prof. Efstathios Kastritis, MD; Plasma Cell Dyscrasia Unit, Amyloidosis Referral Center, Department of Clinical Therapeutics, Alexandra Hospital, Medical School, National and Kapodistrian University of Athens, 11528, 80 Vas. Sofias Str., Athens, Greece. Equal contribution (Raphael Patras, Georgios Georgiopoulos). Equal senior authorship (Efstathios Kastritis, Kimon Stamatelopoulos).

## Abstract

**Background:** Patients with AL amyloidosis present sustained paradoxical vasodilation in response to sympathetic stimulation by cold pressor test (CPT). The clinical relevance of this finding is unknown. We investigated the clinical role of CPT induced vascular and hemodynamic responses.

**Methods:** We prospectively recruited 113 treatment-naïve AL amyloidosis patients. High resolution ultrasonography was used to measure the peak percent change of the brachial artery during CPT and 3 minutes after its’ withdrawal (postCPT), defined as sustained response. Peripheral and aortic (central) systolic (SBP) and diastolic blood pressure (DBP) were measured at the same timepoints before and 12 months after treatment initiation. All-cause and cardiovascular mortality were recorded (median follow-up 26 months). The same tests were performed in ten healthy volunteers.

**Results:** Sustained vasodilation and reductions in central systolic (%CSBP_post) and peripheral diastolic BP were observed in AL as compared to controls (p<0.01 for all) and were associated with all-cause and cardiovascular death after adjustment for disease-related risk factors (p<0.05 for all). %CSBP_post provided incremental value over Mayo stage. Mechanistic analyses revealed associations of %CSBP_post with markers of neurological and cardiac dysfunction and of myocardial infiltration. Longitudinally, at 12 months, %CSBP_post further decreased in patients with earlier poor hematologic response to disease-specific treatment.

**Conclusions:** Using a noninvasive readily available method in treatment-naïve AL amyloidosis patients, sustained reduction of central SBP after sympathetic stimulation was associated with cardiac dysfunction, poor survival and response to treatment.

## Introduction

Light chain amyloidosis (AL) is a potentially lethal systemic disorder caused by the deposition of amyloid fibrils derived from clonal immunoglobulin light chains. Early diagnosis is of paramount importance since the mortality of patients diagnosed with cardiac involvement is high, despite recent improvements in systemic anticlonal therapy^1^. Treatment of AL amyloidosis relies on the elimination of the plasma cell clone, however, adaptations based on the risk of early mortality and the clinical characteristics of the disease are critical to reduce complications and early death rate^2^. Although multiple organs can be affected, cardiac involvement is the main determinant of survival and especially of the risk of early mortality^3^. Vascular involvement, which may be manifested as low blood pressure and paradoxical reactive vasodilation, common manifestations of AL amyloidosis, is also related to poor outcomes independently of heart involvement^4^. Autonomic dysfunction in AL, is considered a major contributor to vascular involvement^4–6^. Although autonomic dysfunction and peripheral nerve involvement, predominantly of small nerve fibers, have been associated with mortality in AL^7^, the clinical role of vascular autonomic dysfunction *per se* has not been explored. More importantly, quantifiable markers for this condition have not been validated for clinical use in AL. We have previously shown that cold pressor test (CPT), an established method to non-invasively induce and investigate vascular responses to sympathetic stimulation, led to paradoxical sustained vasodilation, in a cohort of patients with AL amyloidosis as compared to healthy controls^4^. Interestingly, CPT vasodilation was associated with reactive vasodilation in these patients but not in controls, supporting its’ possible utility as a clinical marker of vascular dysfunction in AL. Based on this evidence, we hypothesized that CPT induced hemodynamic responses could provide a novel clinically meaningful and applicable tool for risk stratification in AL amyloidosis.

## Methods

### Design and population

This is a cohort study in which we prospectively recruited consecutive treatment-naïve patients with systemic AL amyloidosis. All patients were managed and followed in a referral center for amyloidosis in Greece, in the Department of Clinical Therapeutics, Alexandra General Hospital. All subjects were submitted to CPT after detailed history was obtained and thorough clinical examination was performed. All measurements were performed in the Unit of Angiology and Endothelial Pathophysiology, of the center. From January 2019 to December 2022, we consecutively recruited 127 newly diagnosed treatment naïve patients with AL amyloidosis. All subjects were followed for all-cause mortality and cardiovascular (CV) death (co-primary endpoints) for a median of 26 months. Our patient cohort was thoroughly assessed, treated with contemporary regimens and followed closely for outcomes of interest. Consensus criteria were used for the definition and assessment of organ involvement^8^. The research protocol did not interfere with the management of the patients. Criteria for interruption were predefined as numeric rating scale (NRS)>4^9^. Ten patients *a priori* denied undergoing CPT to avoid cold discomfort, while in three patients the test was interrupted due to an NRS>4 and one patient interrupted the test earlier. Hence, 113 patients were included. All patients in the study underwent transthoracic echocardiography at baseline which included both standard echocardiographic imaging and images suitable for 7 speckle tracking analysis, as previously described^10,11^. Further details regarding echocardiography are provided in the **Supplemental Material**. Ten healthy volunteers (controls) without AL amyloidosis or traditional risk factors for cardiovascular disease (mean age 28, male sex 4 (40%)) were also recruited among the staff of our referral center and underwent all CPT tests. The study was conducted according to the Declaration of Helsinki and all patients provided a written informed consent form for anonymous publication of scientific data. The local Institutional Review Board approved the study protocol.

### Cold Pressor Test and Peripheral Vascular Assessment

All CPT parameters and formulas used to calculate them are depicted in **Table 1**. CPT constitutes a reliable method of peripheral sympathetic nervous system stimulation to study its’ effect on hemodynamic parameters and vascular reactivity^12^. Aortic (central) (CSBP) and peripheral (SBP) systolic blood pressure and diastolic blood pressure (CDBP and DBP, respectively) were measured before and 3 minutes after CPT withdrawal. Peripheral blood pressures were additionally measured at the end of CPT stimulus, because measurement of central BP required dedicated equipment not possible to be deployed during the test. Peak percent change of the brachial artery diameter was computed while on CPT (%D_on), that is during the 3 minutes of CPT application, and post CPT (%D_post), that is 3 minutes after CPT withdrawal. PWV was used as the gold-standard non-invasive clinical marker of arterial stiffness (Complior, Art Med, France)^13^ and aortic pressures and markers of arterial wave reflections were estimated using radial artery tonometry (Sphygmocor System-Atcor Sydney, Australia)^14^. On a different visit, all subjects underwent assessment of reactive vasodilation in response to ischemic stimulus using flow-mediated vasodilation (FMD) a well validated non-invasive method for this purpose, as previously described^13^. Brachial diameter measurements were assessed using high resolution ultrasonography (14.0-MHz multifrequency linear array probe, Vivid 7 Pro until February 2021 and 12.0-MHz multifrequency linear array probe, Vivid S70N from March 2021 until the end of recruitment; General Electric Healthcare, Milwaukee, Wisconsin, USA). All hemodynamic tests were also performed 12 months after treatment initiation in all survivors. Further details are provided in **Supplemental Material**.

**Table 1.**
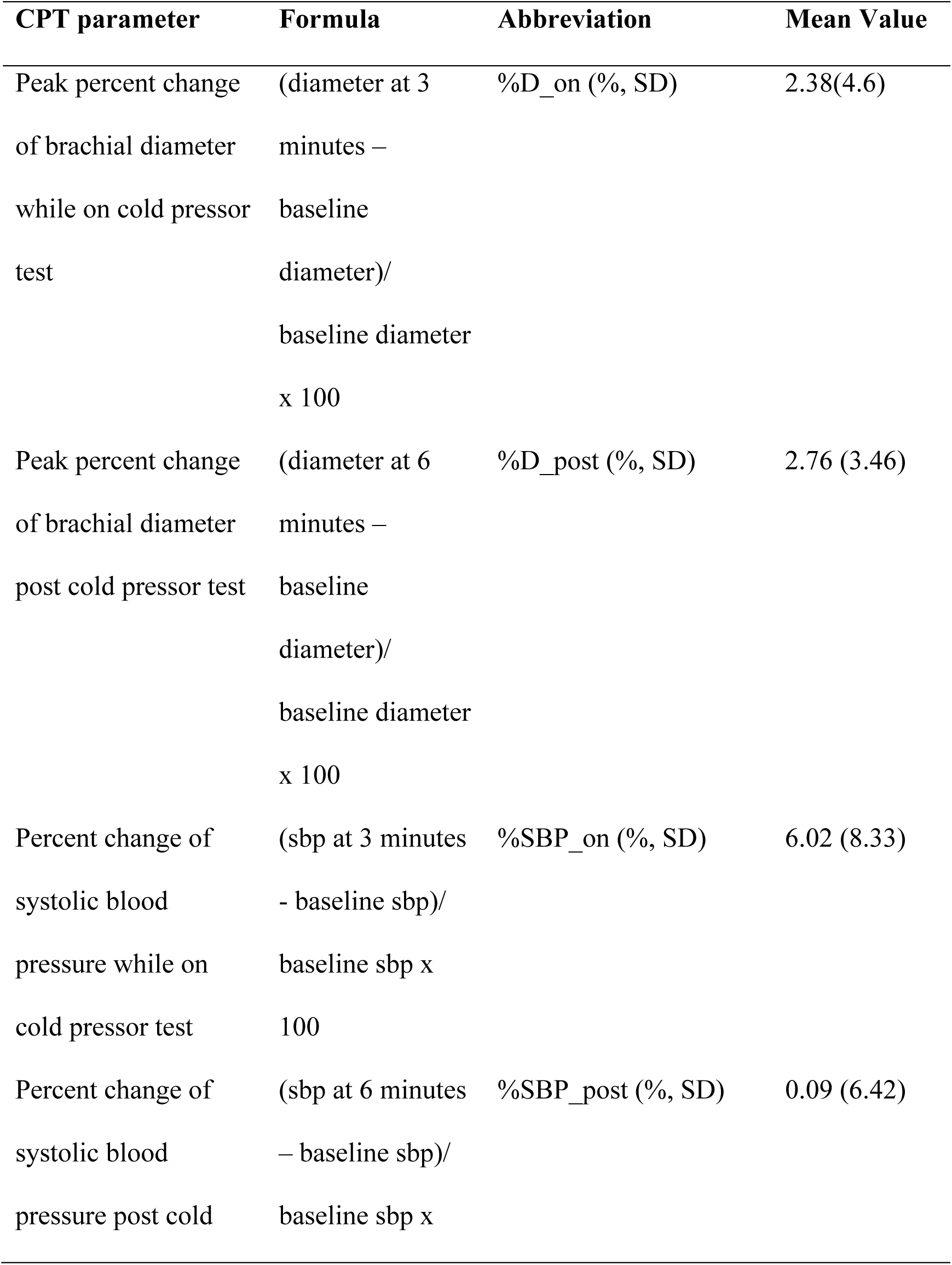
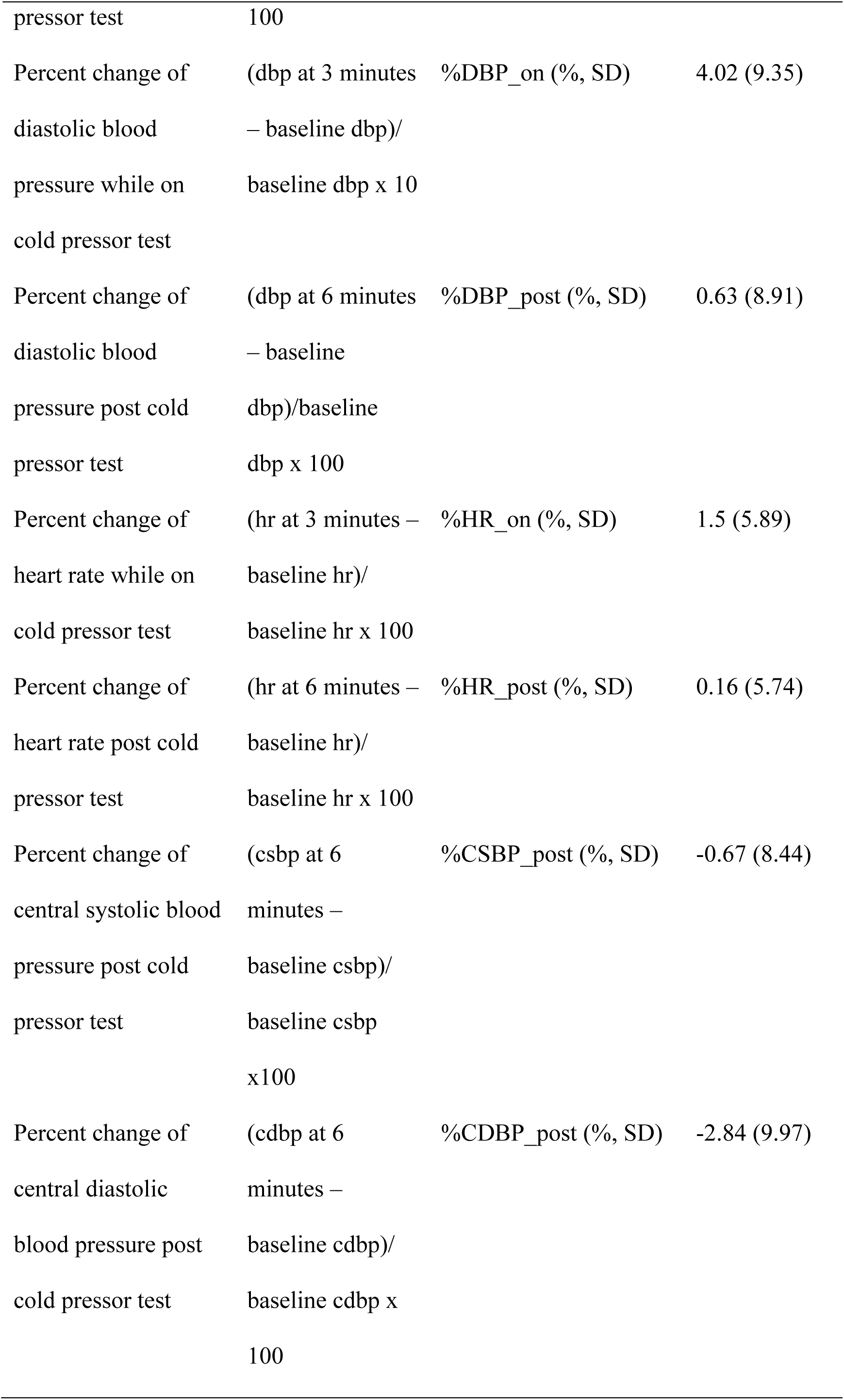
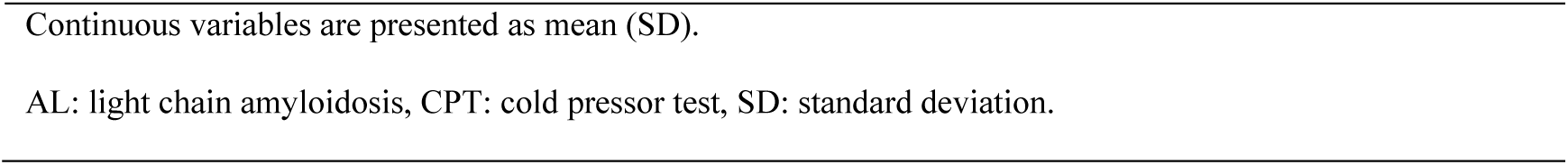
CPT parameters of AL population.

#### Methods used in subgroup analyses

Based on our findings showing the clinical value of hemodynamic responses to CPT, we aimed to further interpret these results by using already available mechanistic information in our cohort. We retrospectively collected results of neurological and cardiac tests performed in our patients with available CPT examination. All tests were eligible if performed during the diagnostic workup of these patients^15^ or under other on-going observational research protocols^7^ within 3 months from index CPT and before treatment initiation. All methods and subgroup descriptive characteristics are outlined in **Supplemental Material** and **Supplemental Table 1.**

In brief, cardiac magnetic resonance (CMR) examination was performed as previously described^15^. Ambulatory blood pressure monitoring (ABPM) was performed on a usual working day. Dippers were defined as subjects with a nocturnal BP decrease > 10% and non-dippers as patients with a nocturnal BP decrease <10%. Baroreceptor reflex sensitivity (BRS) assessment was performed. Chest leads for continuous electrocardiogram recording and a finger cuff of the non-invasive BP monitor (Finometer) were set^16,17^. Sudoscan device evaluates sweat gland innervation at hands and feet by measuring electrochemical skin conductance (ESC) based on a chronoamperometric method and is a useful method for the assessment of autonomic function^18,19^. Sudomotor dysfunction is divided based on ESC values at the level of the feet into two categories: >70μS no dysfunction, <70μS dysfunction^18,20^. Detailed clinical neurological examination using the Neuropathy Symptom Scale (NSS) and Neurologic Impairment Score (NIS) was performed in the first Department of Neurology, National and Kapodistrian University of Athens. Quantitative sensory testing was also performed. Warm detection threshold (WDT), cold detection threshold (CDT), heat pain threshold (HPT) and cold pain threshold (CPT) for the skin of thenar eminence and sole of the foot were evaluated.

## Statistical Methods

Continuous variables are reported as mean ± standard deviation or median and interquartile range for non-normal distributions, while categorical variables are summarized as counts and percentages. Normality was assessed using histograms and Q-Q plots.

We used univariable and multivariable Cox regression models to assess the association between CPT parameters and mortality, reporting hazard ratios (HR) with 95% confidence intervals (CIs). Multivariable models adjusted for age and sex, and proportional hazards assumptions were tested via Schoenfeld residuals. Findings were visualized with Nelson-Aalen curves, and survival functions were compared using the log-rank test. To analyze the prognostic value of CPT-induced hemodynamic and vascular changes, we applied ROC analysis, selecting the optimal cutoff via the Liu method. The incremental value of CPT parameters for mortality prediction in AL amyloidosis was assessed using differences in AUC from time-dependent C-statistics (Harrell’s C) with censored data. AUC comparisons and 95% CIs were computed using the DeLong approach^21^. Statistical analysis was performed in STATA version 18 (StataCorp, USA), with significance set at α=0.05 (two-tailed tests).

## Results

Descriptive characteristics of our AL patient cohort are outlined in **Table 2**. Mean age was 66 years and most patients were males. Heart involvement was present in 83.2%. Most of the patients (59.8%) were classified as NYHA stage 2 or higher at the time of presentation. 45.5% of the cohort was categorized as Mayo stage 3. Renal involvement was present in 57.5% of the patients while clinically detectable involvement of the peripheral nervous system in 21.3%; 8 patients had history of coronary artery disease. Compared to healthy controls, AL patients had higher vasodilation and lower CSBP, SBP and DBP post CPT. (**Supplemental Figure 1**).

**Table 2.**
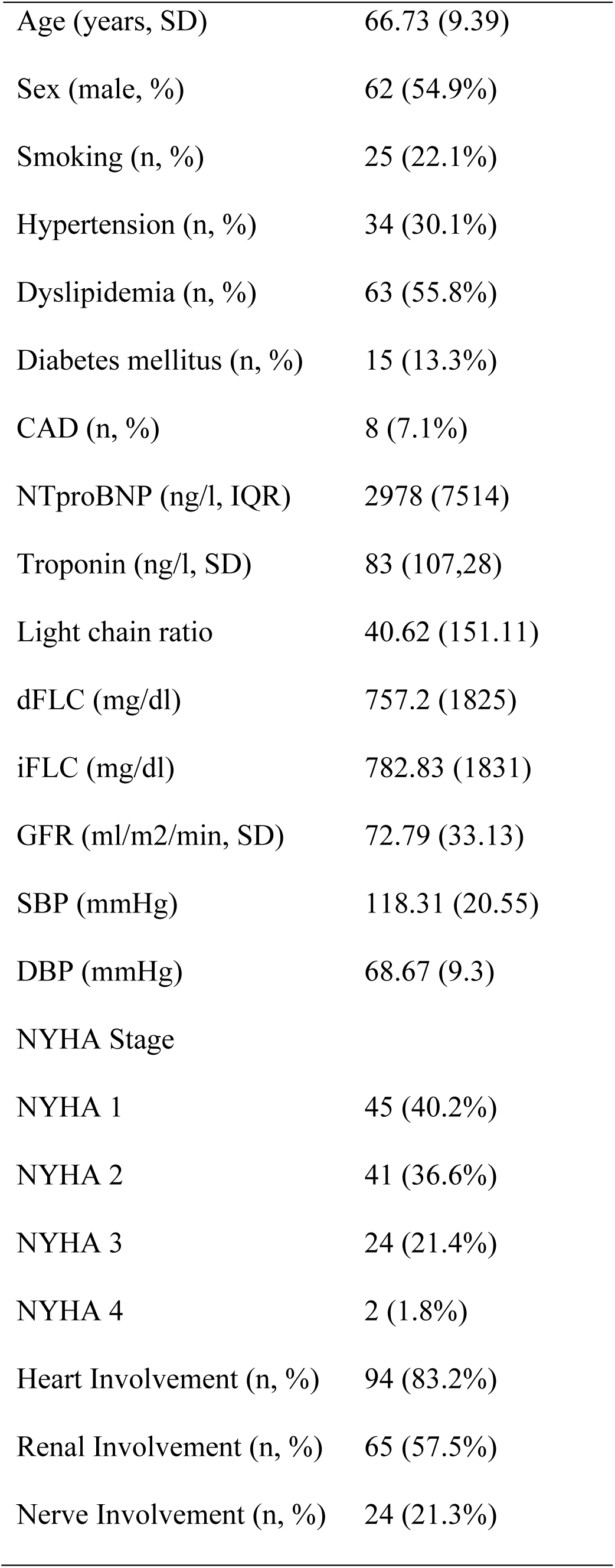
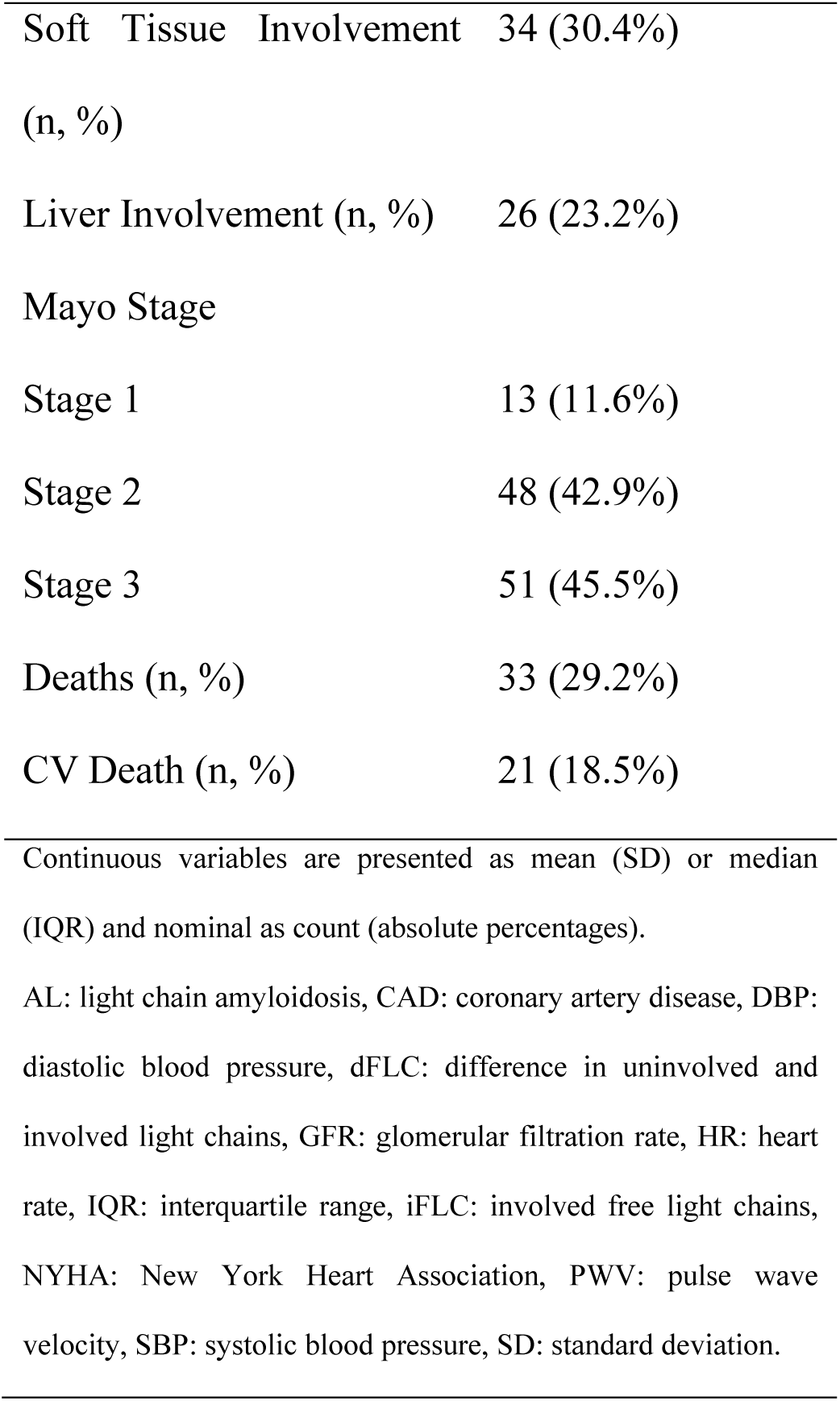
Descriptive characteristics of AL population.

### Prognostic associations of CPT parameters with survival outcomes

After a median follow up period of 26 months, 33 deaths were recorded; 19 of them occurred earlier than 6 months after treatment initiation, while 21 events were of cardiovascular origin. The associations between change in vascular and hemodynamic parameters and mortality endpoints are summarized in **Table 3**.

**Table 3.**
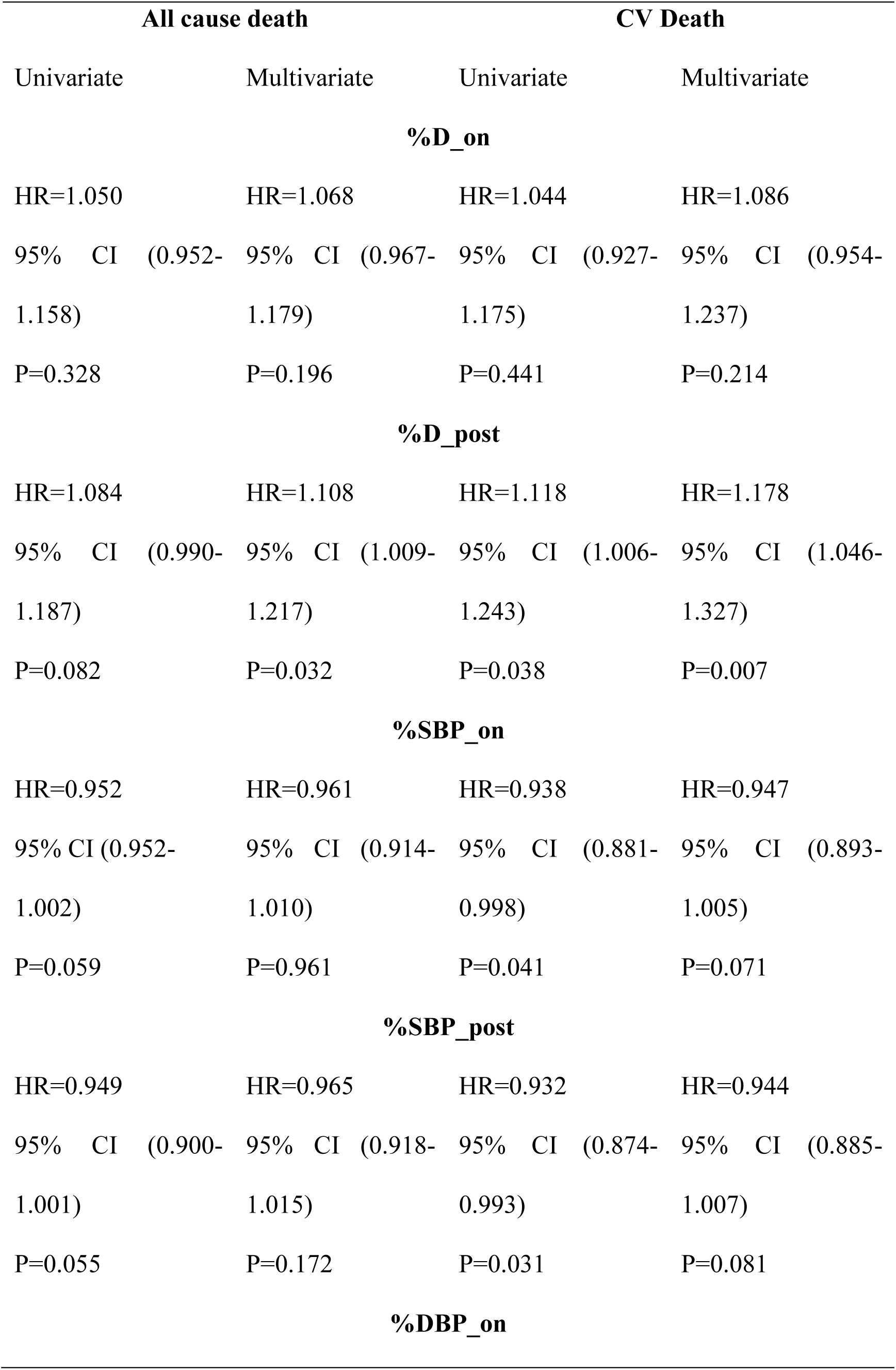
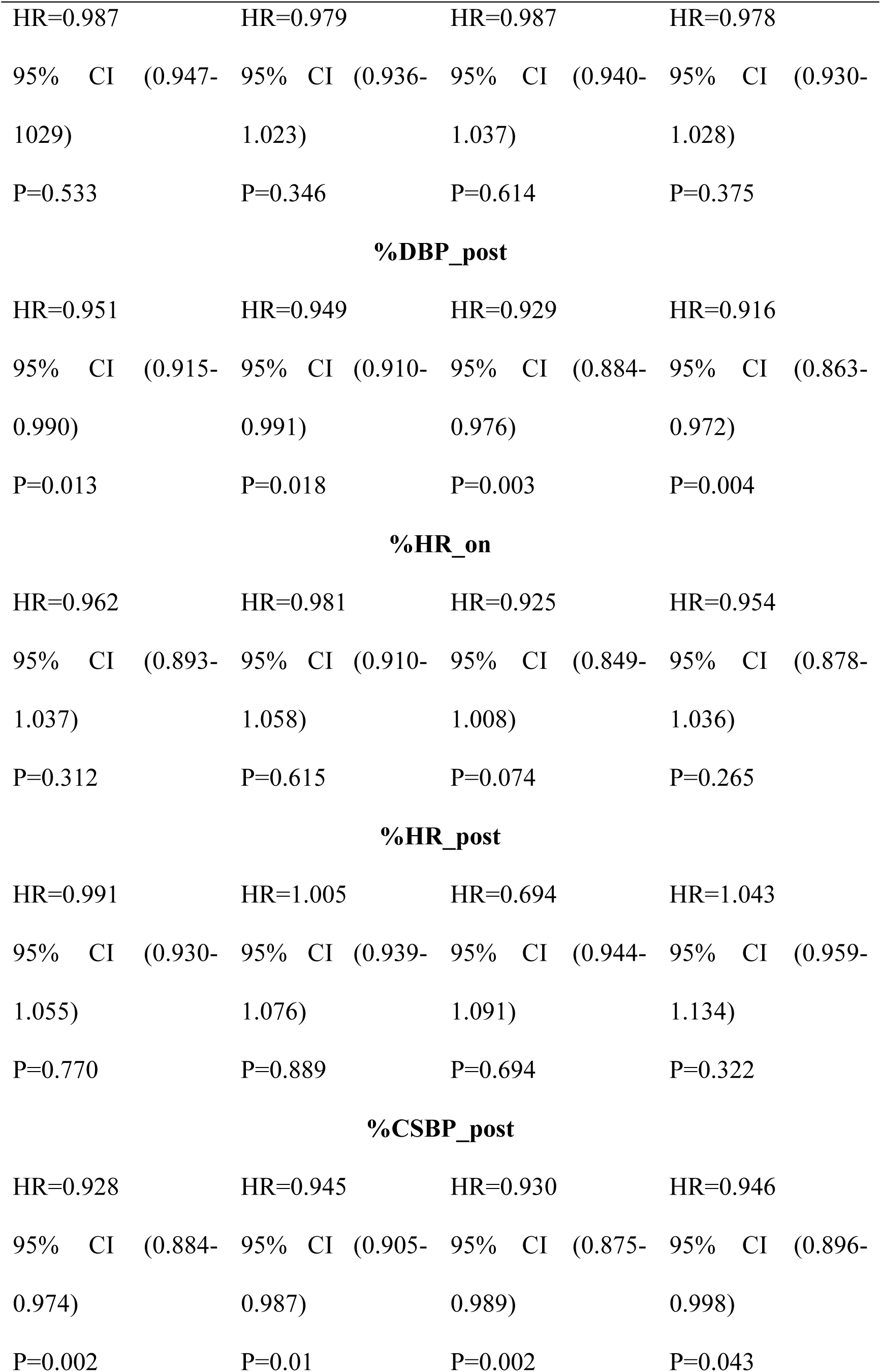
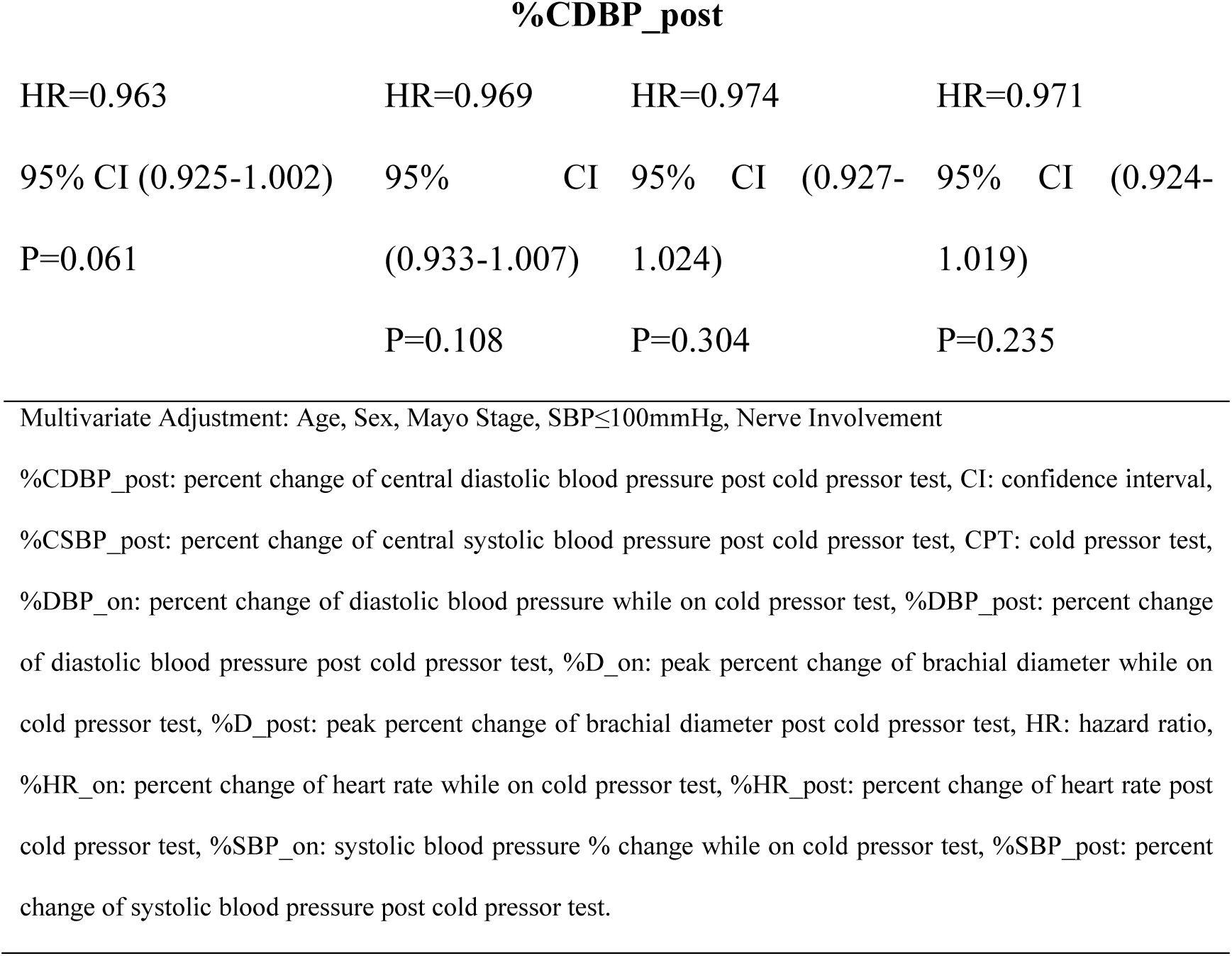
Prognostic Cox models for the association of CPT parameters with mortality.

| All cause death |  |  |  | CV Death |  |  |  |
| --- | --- | --- | --- | --- | --- | --- | --- |
| Univariate |  | Multivariate |  | Univariate |  | Multivariate |  |
| %D_on |  |  |  |  |  |  |  |
| HR=1.050 |  | HR=1.068 |  | HR=1.044 |  | HR=1.086 |  |
| 95% CI | (0.952-1.158) | 95% CI | (0.967-1.179) | 95% CI | (0.927-1.175) | 95% CI | (0.954-1.237) |
| P=0.328 |  | P=0.196 |  | P=0.441 |  | P=0.214 |  |
| %D_post |  |  |  |  |  |  |  |
| HR=1.084 |  | HR=1.108 |  | HR=1.118 |  | HR=1.178 |  |
| 95% CI | (0.990-1.187) | 95% CI | (1.009-1.217) | 95% CI | (1.006-1.243) | 95% CI | (1.046-1.327) |
| P=0.082 |  | P=0.032 |  | P=0.038 |  | P=0.007 |  |
| %SBP_on |  |  |  |  |  |  |  |
| HR=0.952 |  | HR=0.961 |  | HR=0.938 |  | HR=0.947 |  |
| 95% CI | (0.952-1.002) | 95% CI | (0.914-1.010) | 95% CI | (0.881-0.998) | 95% CI | (0.893-1.005) |
| P=0.059 |  | P=0.961 |  | P=0.041 |  | P=0.071 |  |
| %SBP_post |  |  |  |  |  |  |  |
| HR=0.949 |  | HR=0.965 |  | HR=0.932 |  | HR=0.944 |  |
| 95% CI | (0.900-1.001) | 95% CI | (0.918-1.015) | 95% CI | (0.874-0.993) | 95% CI | (0.885-1.007) |
| P=0.055 |  | P=0.172 |  | P=0.031 |  | P=0.081 |  |
| %DBP_on |  |  |  |  |  |  |  |

|  |  |  |  |
| --- | --- | --- | --- |
| HR=0.987 | HR=0.979 | HR=0.987 | HR=0.978 |
| 95% CI (0.947-1.029) | 95% CI (0.936-1.023) | 95% CI (0.940-1.037) | 95% CI (0.930-1.028) |
| P=0.533 | P=0.346 | P=0.614 | P=0.375 |
| **%DBP_post** |  |  |  |
| HR=0.951 | HR=0.949 | HR=0.929 | HR=0.916 |
| 95% CI (0.915-0.990) | 95% CI (0.910-0.991) | 95% CI (0.884-0.976) | 95% CI (0.863-0.972) |
| P=0.013 | P=0.018 | P=0.003 | P=0.004 |
| **%HR_on** |  |  |  |
| HR=0.962 | HR=0.981 | HR=0.925 | HR=0.954 |
| 95% CI (0.893-1.037) | 95% CI (0.910-1.058) | 95% CI (0.849-1.008) | 95% CI (0.878-1.036) |
| P=0.312 | P=0.615 | P=0.074 | P=0.265 |
| **%HR_post** |  |  |  |
| HR=0.991 | HR=1.005 | HR=0.694 | HR=1.043 |
| 95% CI (0.930-1.055) | 95% CI (0.939-1.076) | 95% CI (0.944-1.091) | 95% CI (0.959-1.134) |
| P=0.770 | P=0.889 | P=0.694 | P=0.322 |
| **%CSBP_post** |  |  |  |
| HR=0.928 | HR=0.945 | HR=0.930 | HR=0.946 |
| 95% CI (0.884-0.974) | 95% CI (0.905-0.987) | 95% CI (0.875-0.989) | 95% CI (0.896-0.998) |
| P=0.002 | P=0.01 | P=0.002 | P=0.043 |

**%CDBP\_post**
|  |  |  |  |
| --- | --- | --- | --- |
| HR=0.963 | HR=0.969 | HR=0.974 | HR=0.971 |
| 95% CI (0.925-1.002) | 95% CI (0.933-1.007) | 95% CI (0.927-1.024) | 95% CI (0.924-1.019) |
| P=0.061 | P=0.108 | P=0.304 | P=0.235 |

Multivariate Adjustment: Age, Sex, Mayo Stage, SBP≤100mmHg, Nerve Involvement
%CDBP\_post: percent change of central diastolic blood pressure post cold pressor test, CI: confidence interval, %CSBP\_post: percent change of central systolic blood pressure post cold pressor test, CPT: cold pressor test, %DBP\_on: percent change of diastolic blood pressure while on cold pressor test, %DBP\_post: percent change of diastolic blood pressure post cold pressor test, %D\_on: peak percent change of brachial diameter while on cold pressor test, %D\_post: peak percent change of brachial diameter post cold pressor test, HR: hazard ratio, %HR\_on: percent change of heart rate while on cold pressor test, %HR\_post: percent change of heart rate post cold pressor test, %SBP\_on: systolic blood pressure % change while on cold pressor test, %SBP\_post: percent change of systolic blood pressure post cold pressor test.

After multivariable adjustment, %CSBP_post, %D_post and %DBP_post were independent predictors of mortality (all cause death: HR=1.108 95% CI (1.009-1.217) p=0.032, HR=0.949 95% CI (0.910-0.991) p=0.018, HR=0.945 95% CI (0.905-0.987) p=0.01 respectively, CV death: HR=1.178 95% CI (1.046-1.327) p=0.007, HR=0.916 95% CI (0.863-0.972) p=0.004, HR=0.946 95% CI (0.896-0.998) p=0.043 respectively) (**Table 3**).

**Figure 1** depicts Kaplan Meier survival curves for all-cause death and CV death. Survival was worse in patients with a prominent ROC-derived CSBP reduction>3.23% (log rank test, p=0.011 for all-cause death and p=0.027 for CV death) and DBP reduction>4.23% (log rank test, p=0.033 for all-cause death and p=0.05 for CV death). Regarding %D_post, we found that a prominent ROC-derived increase>5.88% was associated with increased risk of all-cause and CV death (log rank test, p=0.001 for both).

**Figure 1.**
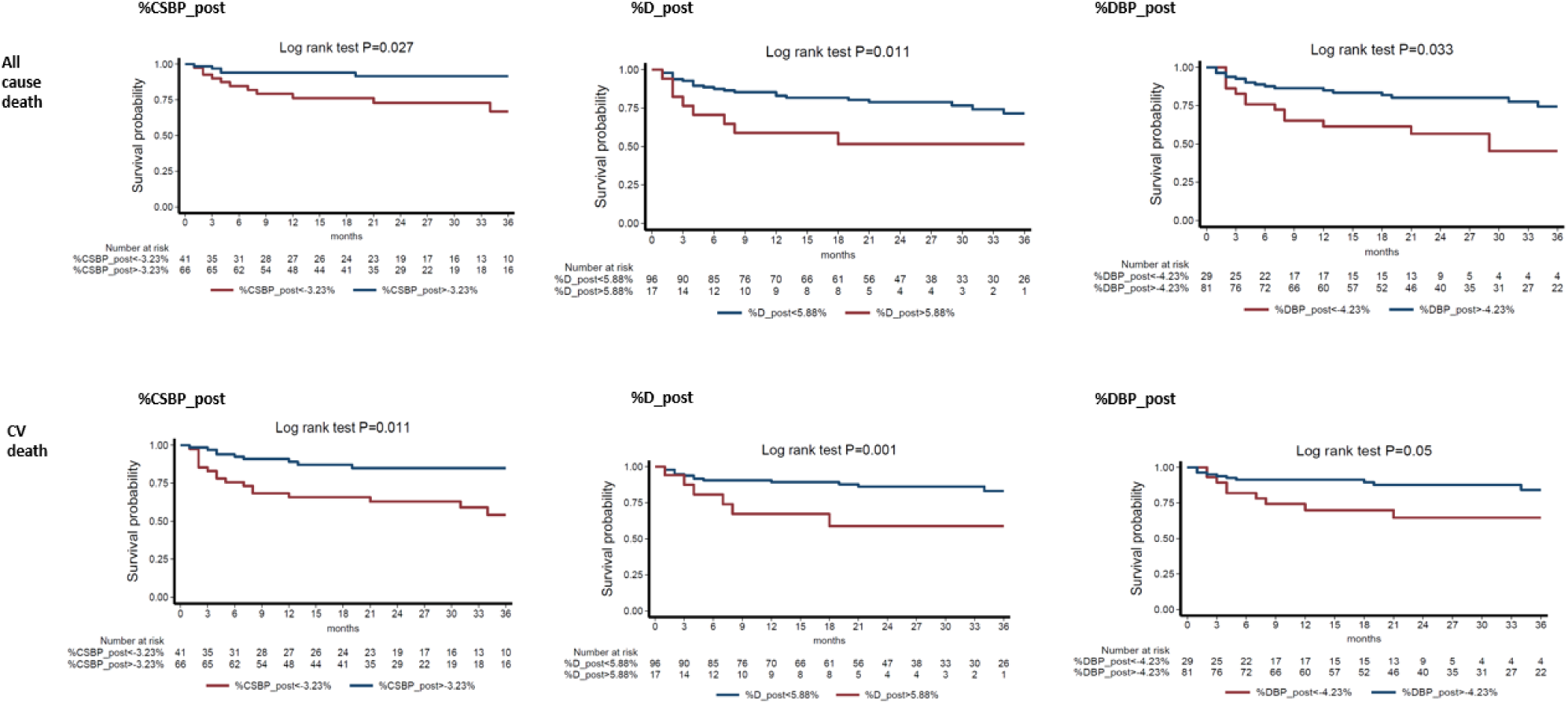
Kaplan-Meier survival curves for all-cause and CV death. Patients with a decrease in %CSBP_post more than 3.23% and in %DBP_post more than 4.23% had worse survival. %D-post >5.88% was related to increased risk of death. %CSBP_post: percent change of central systolic blood pressure post cold pressor test, %DBP_post: percent change of diastolic blood pressure post cold pressor test, %D_post: peak percent change of brachial diameter post cold pressor test.

There were significant correlations between CPT parameters and markers of disease severity (**Supplemental Table 2**). Briefly, %D_post was significantly associated with NTproBNP levels and symptomatic heart failure by NYHA stage, reflecting cardiac involvement (p for all <0.05). Both %CSBP_post and %SBP_post displayed an inverse correlation with Mayo stage, reflecting disease severity and poor prognosis (p for all <0.05).

Of all CPT parameters, %CSBP_post provided incremental value over the most widely used prognostic score for AL amyloidosis, the Mayo stage^22^. In specific, %CSBP_post conferred incremental prognostic value over the Mayo stage for all-cause (Mayo stage AUC 0.685 95% CI 0.604-0.764, Mayo stage + %CSBP_post AUC 0.748 95% CI (0.650-0.845), p=0.047) and CV mortality (Mayo stage AUC 0.749 95% CI 0.673-0.824, Mayo stage + %CSBP_post AUC 0.806 95% CI 0.723-0.889, p=0.046).

### Hematologic response predicts changes in CPT parameters

The main goal of treatment in AL amyloidosis is the elimination of the production of toxic free light chains, and the depth of hematologic response has been associated with organ function recovery and improved survival. Subjects with no hematologic response (NR) or those that had only achieved a partial hematologic response (PR) at 6 months after treatment initiation displayed a decrease in %CSBP_post at 12 months (p=0.047); in contrast, those with complete hematologic (CR) or very good partial hematologic response (VGPR) at 6 months exhibited a nonsignificant increase in %CSBP_post at 12 months, resulting in a significant difference between the two groups (p for interaction=0.003) (**Figure 2**). These results indicate that poor hematologic response is associated with further deterioration of central blood pressure probably because of continuous toxic immunoglobulin light chain and amyloid production.

**Figure 2.**
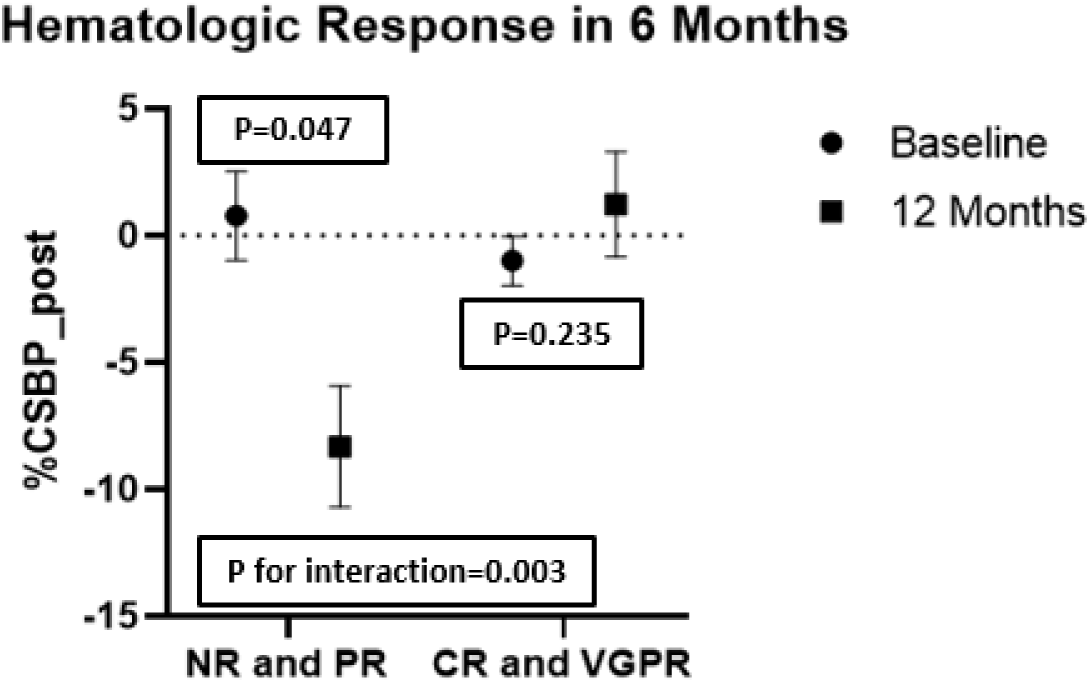
Change of %CSBP_post after treatment initiation. Patients with NR or PR 6 months after treatment initiation exhibited a decrease in %CSBP_post while those with CR or VGPR displayed an increase, in 12 months. CPT: cold pressor test, CR: complete response, %CSBP_post: percent change of central systolic blood pressure post cold pressor test, NR: no response, PR: partial response, VGPR: very good partial response.

### Analysis for mechanistic associations

Associations with echocardiographic markers are presented in **Supplemental Table 3.** %CSBP_post exhibited a negative association with LV wall thickness, left atrium (LA) diameter, global longitudinal strain (GLS) and a positive relationship with LVEF (p for all <0.05). %SBP_post was positively related to spectral tissue doppler imaging (STDI) (p=0.029). %DBP_post displayed a positive relationship with left ventricular ejection fraction (LVEF) and an inverse one with E/e’, a well-known marker of diastolic dysfunction (p for all <0.05). %D_post presented positive correlation with FMD (p=0.002). No other correlation between vascular function markers and CPT parameters was revealed (**Supplemental Table 4).**

### Subgroup Mechanistic Analyses

Associations between CPT parameters and markers of neurologic assessment are depicted in **Supplemental Table 5**. Overall, lower %CSBP was most prominently and consistently associated with markers of neurologic and cardiac dysfunction. In specific, %CSBP_post correlated positively with higher heart rate variation assessed by (coefficient of variability of heart rate), CDT of the hand and negatively with HDT and HPT of the hand reflecting involvement of the autonomic component of the nervous system (p for all <0.05). Additionally, %CSBP_post exhibited negative relationships with the sensory and total score of NSS as well as motor, sensory and total score of NIS (p for all <0.05). Subjects with CDT of the hand <29°C and HDT of the hand>44°C exhibited lower %CSBP_post as opposed to their counterparts (**Figure 3A-B**). Finally, in subjects with dysfunction of the sensory component of the nervous system, as assessed by NISS, %CSBP_post was lower (**Figure 3C**). Of all CPT parameters, only %CSBP_post displayed significant correlations with CMR markers, such as T1, T2, extracellular volume (ECV) and late gadolinium enhancement (LGE) (p for all <0.05) (**Supplemental Table 6**). Lower %CSBP_post was associated with higher GLS and T1, T2 (**Figure 3D-F)**. Among the rest of CPT markers, %D_post was more profound in non-dippers than dippers (**Supplemental Figure 2A**). Most subjects with sudomotor dysfunction (ESC<70μS) had %D_post>5.88% (**Supplemental Figure 2B**). %DBP_post was lower in those with autonomic neuropathy as assessed by NSSA (**Supplemental Figure 2C**). Taken together, these findings suggest that sustained decreases of central blood pressure in response to a sympathetic stimulus may be linked with deteriorated cardiac systolic function and adverse myocardial tissue remodeling as well as with markers of neurological dysfunction.

**Figure 3.**
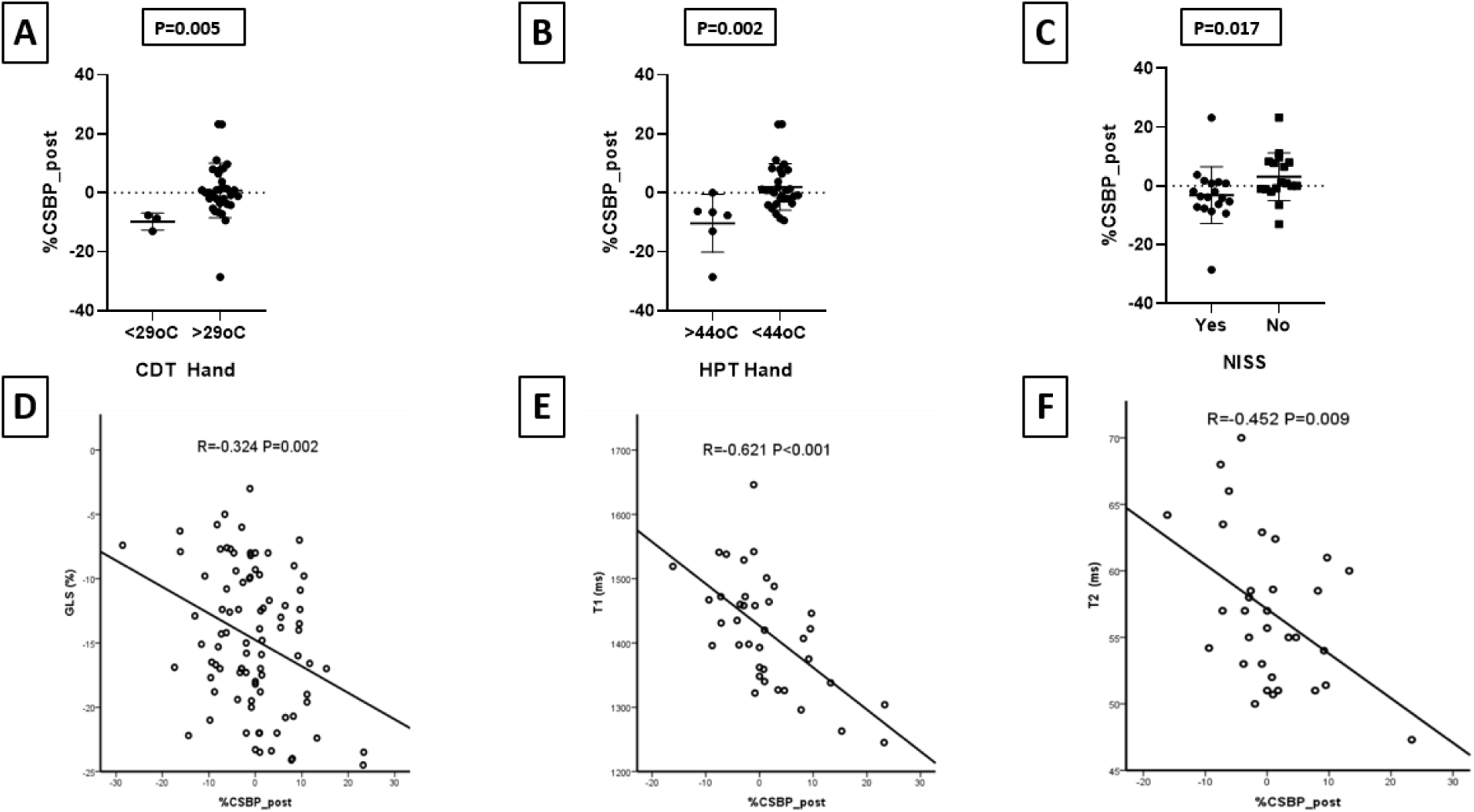
Association between %CSBP_post and markers of neurologic and cardiac function. %CSBP_post was lower in those with CDT of the hand <29°C, HPT of the hand>44°C and neurologic dysfunction assessed by NISS. %CSBP_post displayed a negative correlation with T1 and T2 in CMR. In addition, %CSBP_post was negatively correlated with GLS. CDT: cold detection threshold, CMR: cardiac magnetic resonance, %CSBP_post: percent change of central systolic blood pressure post cold pressor test, GLS: global longitudinal strain, HPT: heat pain threshold, NISS: neurologic impairment score sensory, NSSA: neuropathy symptom scale autonomous subscore

## Discussion

The current study investigates for the first time the clinical utility of hemodynamic responses to sympathetic stimulation in AL amyloidosis, using a readily applicable non-invasive methodology. We found that paradoxical reduction of blood pressure and sustained vasodilation of the brachial artery, induced by CPT, were associated with all-cause and CV mortality. Particularly, sustained reduction of CSBP, which is readily measured by increasingly available dedicated devices, provided the most consistent and clinically meaningful associations with markers of cardiac and neurologic dysfunction providing incremental value over the Mayo staging system. Importantly, poor hematologic response to anti-clonal therapy was associated with further deterioration of %CSBP_post. These findings pave the way for further assessment of the clinical utility of %CSBP_post as a prognostic biomarker in AL amyloidosis (**Graphical Abstract**).

The contribution of vascular involvement to risk stratification in AL patients is still under investigation. We have previously demonstrated that reactive vasodilation of the brachial artery is increased in AL patients and is associated with increased mortality with incremental value over heart involvement and hypotension^4^. Increased FMD was closely associated with vascular autonomic dysfunction, manifested as paradoxical vasodilation in response to CPT^4^. In contrast, controls without amyloidosis as well as the general population and patients with cardiovascular disease (CVD) risk factors or established CVD typically manifest vasoconstriction or no response^4,12^. These results, combined with the commonly found autonomic dysfunction in AL patients^7,23^, support the hypothesis that vascular autonomic dysfunction may be one of the hallmarks of vascular involvement in AL amyloidosis. To further explore whether vascular dysfunction is incrementally driving increased mortality in AL amyloidosis beyond heart involvement, we comprehensively assessed the clinical relevance of hemodynamic responses to CPT, in a population of amyloidosis patients with available survival and mechanistic data. Our findings further support the clinical utility of markers of vascular dysfunction, specifically highlighting the decrease of CSBP after CPT in AL amyloidosis. Specifically, CSBP changes after CPT withdrawal were consistently associated with multiple structural and functional cardiac markers and CMR derived myocardial tissue indices characteristic for amyloidosis, suggesting that this marker may cumulatively reflect cardiac involvement and its’ severity. As compared to peripheral SBP, aortic blood pressure informs more accurately on the interaction of arterial wave reflections with cardiac output and therefore on left ventricular afterload^24^. These attributes may explain our observations, suggesting a clinically meaningful role of CSBP response to CPT as a marker of arterial-ventricular coupling in AL amyloidosis. To that end, we found that CSBP drop after CPT was further deteriorated in patients who presented poor hematologic response, which is consistent with recently described improvements of markers of cardiac involvement in AL patients according to treatment response^11^. Therefore, %CSBP_post may be a clinically relevant organ-specific therapeutic biomarker in AL amyloidosis, warranting further research. On the other hand, while sustained vasodilation after CPT was only weakly associated with cardiac involvement it was the only parameter that correlated with FMD of the brachial artery, confirming our previous findings^4^ . This implies that vasodilation after CPT may act as a specific marker for peripheral vascular involvement in amyloidosis. Collectively, to the best of our knowledge, such patterns of change in vascular and hemodynamic markers have not been previously described for other diseases, suggesting a unique pattern in AL amyloidosis. Given that timely diagnosis and treatment directly affect survival in AL amyloidosis^25^ further research to validate such a specific vascular pattern is warranted.

Current knowledge in the field cannot fully discern the mechanisms leading to sustained paradoxical vasodilation and BP drop in response to sympathetic stress in AL amyloidosis. Our mechanistic findings strongly suggest that autonomic dysfunction may at least partly drive this vascular pattern. Specifically, we found that patients with reduction in their CSBP after CPT, presented lower perception threshold to cold, and higher to heat stimulus and higher pain threshold to heat exposure. Previously, maximum pain perception has been associated with a sharp rise in blood pressure during CPT via sympathetic excitation^26^. Although decreased perception of cold could reduce discomfort leading to a lower rise of blood pressure, a decrease of BP from baseline is unlikely. To that end, in patients with higher reduction in aortic SBP after CPT, we found supportive evidence of global peripheral neurological impairment with more prominent sensory and motor dysfunction and of higher variation of heart rate, a marker of cardiac autonomic dysfunction^27^. In support, in AL amyloidosis, myelinated Αδ fibers and unmyelinated C fibers, which convey thermoregulatory and autonomic information respectively, are primarily affected^18^. On the other hand, sustained vasodilation was associated with non-dipping status, a condition linked with increased prevalence of autonomic neuropathy and orthostatic hypotension among various populations^28,29^. Similarly, it is also associated with higher prevalence of sudomotor dysfunction, a marker of autonomic dysfunction in populations including transthyretin and AL amyloidosis ^18,30,31^.

## Study Limitations

Certain limitations of our study should be acknowledged. Our cross-sectional design demonstrating mechanistic associations of CPT responses to neurological and cardiovascular imaging markers cannot infer causality. Importantly, central blood pressure calculation can be readily measured by widely available devices at relatively low cost and independently of the operator^13^. Therefore, from a methodological point of view, this renders the measurement of %CSBP_post, a readily applicable marker in clinical practice^4,13^.

In conclusion, in patients with AL amyloidosis, paradoxical vasodilation and blood pressure decrease in response to CPT were independently associated with poor outcomes, neurologic dysfunction and more severe cardiac involvement, supporting their utility as markers of disease-related vascular dysfunction. Reduction of CSBP after CPT was the most clinically relevant marker with incremental value over Mayo stage. Given the link we observed between poor hematologic response and further deterioration of aortic blood pressure reduction after sympathetic stimulus, as well as the potentially wide applicability of our methodology, this hemodynamic marker merits further investigation for validation as a novel prognostic and therapeutic biomarker in AL amyloidosis.

## Perspectives

Sustained reduction of arterial blood pressure and vasodilation in response to sympathetic stimulation were associated with worse survival. Poor hematologic response is associated with further deterioration of CSBP after sympathetic stimulation.

## Novelty and Relevance

1. What is new: Novel treatments in AL amyloidosis have improved survival even in patients at ultra-high risk. Previously validated biomarkers may not be as accurate with novel therapies, especially among patients at intermediate- or high-risk.
2. What is relevant: Sustained reduction in aortic or peripheral arterial BP in response to sympathetic stimulation, independently associated with increased mortality, characteristics of advanced disease and worse treatment response.
3. Clinical Implications: Sustained paradoxical reduction in central systolic blood pressure after sympathetic stimulation was consistently associated with all study outcomes, laying the foundations for validation as a prognostic and therapeutic biomarker in AL amyloidosis.

## Data Availability

All data referred in the manuscript are available

## Acknowledgements

nothing to acknowledge.

## Abbreviations List

CPT: cold pressor test
%CSBP_post: percent change of central systolic blood pressure post cold pressor test
%D_post: peak percent change of brachial diameter post cold pressor test
%DBP_post: percent change of diastolic blood pressure post cold pressor test
Light chain amyloidosis: AL amyloidosis

---

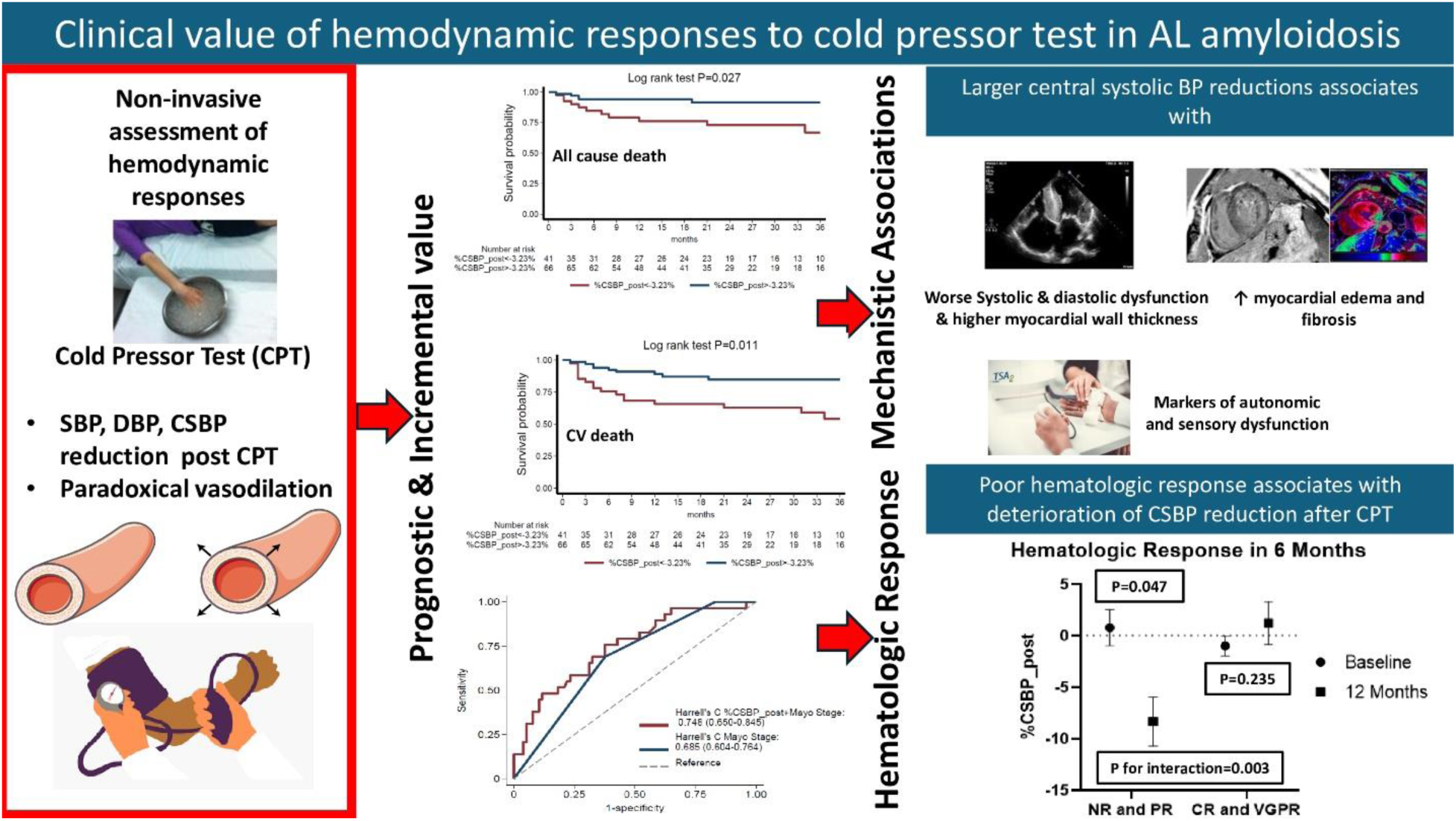

